# Prevalence and predictors of coronaphobia among frontline hospital and public health nurses

**DOI:** 10.1101/2020.10.18.20214692

**Authors:** Leodoro J. Labrague, Janet Alexis A. De Los Santos

## Abstract

**Objectives:** To determine the prevalence rate as well as the predictors of coronaphobia in frontline hospital and public health nurses.

**Design:** This study used a cross-sectional research study involving 736 nurses working in COVID-19 designated hospitals and health units in Region 8, Philippines. Four structured self-report scales were used, including the Coronavirus Anxiety Scale, the Brief Resilience Scale, the Perceived Social Support Questionnaire, and the single-item measure for perceived health.

**Results:** The prevalence rate of coronaphobia was 54.76% (*n* = 402): 37.04% (*n* = 130) in hospital nurses and 70.91% (*n* = 273) in public health nurses. Additionally, nurses’ gender (*β* = 0.146, *p* = 0.001), marital status (*β* = 0.101, *p* = 0.009), job status (*β* = 0.132, *p* = 0.001), and personal resilience (*β* = −0.154, *p* = 0.005) were identified as predictors of COVID-19 anxiety. A small proportion of nurses was willing (19.94%, *n* = 70) and fully prepared (9.40%, *n* = 33) to manage and care for coronavirus patients.

**Conclusion:** Coronaphobia is prevalent among frontline Filipino nurses, particularly among public health nurses. Interventions to address coronaphobia among frontline nurses in the hospital and community should consider the predictors identified. By increasing personal resilience in nurses through theoretically-driven intervention, coronaphobia may be alleviated.

## Introduction

The COVID-19 pandemic is undoubtedly one of the most important challenges to many healthcare systems worldwide. Since the onset of the pandemic, many nations’ healthcare systems, particularly those in low-income countries, have struggled from shortage of manpower, scarcity of supplies and equipment, and other logistics necessary to effectively fight this deadly infection (Hopman *et al*., 2020; McMahon *et al*., 2020). The nursing sectors, being on the frontlines, have been heavily affected by the wrath of the coronavirus disease. Due to the influx of infected patients to the hospitals, many of these nurses have to deal with their fears of the new virus which were amplified due to increased workloads and added responsibilities, new protocols related to COVID-19, and increased nurse-patient ratio, along with the lack of hospital supplies and equipment to effectively manage patients infected with the virus (Al Reshidi, 2020; Lucchini *et al*., 2020). This scenario has caused tremendous challenges to nurses, posing threat to their mental, psychological, and emotional well-being and overall health (Stelnicki *et al*., 2020; Leng *et al*., 2020).

Defined as a “disproportionate state of anxiety or persistent or uncontrollable fear that interferes with daily life and causes disruptions to behaviour and psychological well□being due to the coronavirus pandemic” (Lee *et al*., 2020), coronaphobia, or dysfunctional levels of anxiety related to coronavirus, has been reported by a significant proportion of healthcare workers (HCWs), including nurses. Mounting evidence has shown that approximately 22% to 37% of HCWs suffered from abnormal levels of anxiety due to the virus (Gupta *et al*., 2020; Pappa *et al*. 2020), and among this group of HWCs, nurses reported the highest incidence, which ranged from 25.5% to 92% (Chorwe-Sungani 2020; Saricam, 2020; Labrague & De los Santos, 2020a). Female nurses (Bitan *et al*., 2020; Pappa *et al*., 2020), and those who were assigned in COVID-19 designated institutions, were twice as anxious as those nurses who worked in non-coronavirus specific hospitals (Mora-Magana *et al*., 2020; Pouralizadeh *et al*., 2020). In addition, nurses who perceived inadequate social and workplace support, personal resilience, and coping behaviours were seen to experience elevated levels of anxiety associated with COVID-19 (Cooper *et al*., 2020; Labrague & De los Santos, 2020; Barzilay *et al*., 2020).

Anxiety in frontline nurses mostly originates from their fear of being infected or infecting others, including their family, friends, and peers (Saricam, 2020). Other sources cited by nurses that cause them to be anxious were: insufficient supply of personal protective equipment (PPE), fear of bringing the virus home, lack of access to coronavirus testing, uncertainty that their organisation would sufficiently support them should they be infected, fear of passing on the virus to other healthcare team members, problems related to the availability of childcare services, fear of being assigned to an unfamiliar unit, and lack of access to updated information related to COVID-19 (Shanafelt *et al*., 2020; Wu *et al*., 2009). As higher levels of anxiety are attributed to undesirable outcomes in nurses, including mental and psychological issues, physiological impairment, poor work performance, and low job satisfaction (Labrague & De los Santos, 2020b; Irshad *et al*., 2020), organisational measures must be in place to ensure that these groups of healthcare professionals remain engaged.

As with the other countries, healthcare workers in the Philippines, including nurses, are on the frontlines in the fight against this fatal virus, which has already infected at least 300,000 Filipino and claimed 4,000 lives (Department of Health, 2020; World Health Organization, 2020). This is despite the intensive measures taken by the Philippine government to enhance the general awareness of the community regarding the disease, improve their preventive behaviours to protect themselves from getting the disease, and contain the disease transmission through home confinement measures, social distancing, and strict quarantine protocols. Additionally, in an effort to prevent or curb the spread of the virus and flatten the curve, the Philippine health agency has further strengthened the capacity of the public health units in every area of the country to assist in the overall management of coronavirus cases (Department of Health, 2020). Since the onset of the pandemic, public health nurses actively engaged in the prevention of the spread of coronavirus infection through case surveillance, monitoring of suspected cases, and management of the asymptomatic cases or cases with mild symptoms who were isolated in the designated community isolation facilities.

Given the vital roles frontline hospital and public health nurses play during this pandemic, it is imperative to assess their mental well-being, particularly for the presence of dysfunctional anxiety or coronaphobia, a condition which could adversely affect their work performance, job satisfaction, and overall health and which could possibly drive them to leave their jobs (Labrague & De los Santos, 2020; Stelnicki *et al*., 2020; Leng *et al*., 2020). However, to date, most of the studies focused on mental health outcomes of pandemic in frontline hospital nurses (Chorwe-Sungani 2020; Saricam, 2020), while studies assessing mental and psychological consequences of the pandemic on public health nurses has largely been ignored. Therefore, this study was undertaken to compare the prevalence rate as well as predictors of dysfunctional anxiety between frontline hospital and public health nurses. Findings of this study are essential to guide policymakers in the formulation of strategies to enhance mental health and well-being among frontline nurses so they can effectively perform their role during this coronavirus pandemic.

## Methods

### Research Design

This is a cross-sectional research study using four structured self-report scales.

### Samples and Settings

This study involved frontline hospital and public health nurses in Western Samar, Philippines, from 15 hospitals and 10 health units, which were chosen randomly from the list of all health centres and hospitals within the Region. The sample size requirement was determined using power estimates for 11 predictive variables in multiple linear regression. To achieve an 80% power with a small effect size (0.03) and an alpha set at 0.05, the required sample was 586 nurses as calculated by the G Power program (Soper, 2020). Seven hundred fifty nurses were invited initially, and only 736 nurses responded (351 hospital nurses and 385 public health nurses). Online survey using the Google form was used to gather data from nurses from the selected hospitals and public health units. Nurses with at least six months of working experience in the hospital and health centres were included in the study.

### Instrumentation

Five standardized questionnaires were used in this study including the Coronavirus Anxiety Scale (CAS) (Lee *et al*., 2020), the Brief Resilience Scale (BRS) (Smith *et al*., 2008), Perceived Social Support Questionnaire (PSSQ) (Lin *et al*., 2019), and the single-item measure for perceived health.

The 5-item CAS was the primary instrument used to determine coronaphobia. Nurses responded through a five-point Likert scale ranging from “not at all” to “nearly every day”. To discriminate coronaphobia from normal anxiety, the cut off score of 9 set by the original author (Lee *et al*., 2020) was followed. The scale had excellent psychometric properties and had previously successfully validated for use in healthcare workers (Mora-Magana *et al*., 2020). In this study, the internal consistency value was 0.87.

The BRS was used to assess nurses’ ability to bounce back from traumatic or distressing situations due to pandemic. The scale was completed by nurses by responding to each item using a 5□point Likert□type scale running from “does not describe me at all” to “describes me very well”. The validity and reliability of the scale were found to be optimal based on previous studies (Smith *et al*., 2008) and an acceptable internal consistency value of 0.84 in this research.

The PSSQ was used to examine the degree of social support nurses’ received during the coronavirus pandemic. Nurses completed the scale by responding to each item on the 5□point Likert□type scale ranging from “strongly disagree” to “strongly agree”. The validity and the reliability of the scale were previously established with an acceptable internal consistency of 0.89 (Lin *et al*., 2019) and internal consistency value of 0.90 in this research.

A single-item questionnaire was used for the overall assessment of nurses’ health. Nurses rated their overall health using a 5-point Likert scale ranging from “poor” to “excellent”. Previous research established the reliability of the item with a test-retest reliability value of 0.91 (Labrague *et al*., 2020a) and a reliability value of 0.88 in the current research.

### Data Collection and Ethical Considerations

The research proposal was sent to the Institutional Research Ethics Committee of Samar State University (IRERC EA□0012□I). After the approval of the study, the online survey questionnaire using a Google form was sent to email addresses of all prospective nurses within the region. The front page of the Google form contained the basic information regarding the research as well as the letter of consent. To ensure the anonymity of the participants, no background information was needed to fill the online survey. The survey was conducted for a duration of one month from 1 September to 1 October 2020, which coincides with the sixth month of the mandatory lockdown due to the COVID-19 pandemic. Weekly reminders through email were sent to nurses to follow up and remind them to complete the survey.

### Data Analysis

Descriptive and inferential statistics were used to analyse the data gathered using the SPSS version 25. Descriptive data were analysed using the frequencies, standard deviations, and means. Relationships between students’ variables and key study variables were examined using the independent *t*-test, Pearson’s *r* correlation coefficient, and analysis of variance (ANOVA). Using multiple linear regression, variables which significantly correlated with the outcome variable through the bivariate analyses were interred into the model to identify possible predictors of lockdown fatigue. Prior to the regression analysis, multicollinearity was examined by correlating the key study variables.

## Results

Seven hundred one nurses were recruited to partake in the study: 386 public health nurses and 325 hospital nurses. The mean age of nurses was 31.87 years (SD: 7.35). The average years of experience in nursing and in their organisation was 8.39 years and 5.16 years, respectively. Most of the participants were female (78.10%) and held BSN degrees (87.89%) and full-time job status (79.32%). Half of the participants (50.75%) had no attendance to any training related to coronavirus, while almost all of them (95.24%) were aware of the presence of COVID-19 protocols in their workplace. Only 19.94% perceived themselves as “absolutely willing” to manage infected patients, while less than 10% perceived themselves as “fully prepared” to care for coronavirus patients **(Table 1)**.

**Table 1.** Nurse Characteristics (*n* = 736)

| Characteristics | Category | Hospital Nurses<br>N (%) (*n* = 351) | Public Health<br>Nurses<br>N (%) (*n* = 385) | All Nurses<br>N (%) (*n* = 736) |
| --- | --- | --- | --- | --- |
| Age |  | 30.941 (6.769) | 32.654 (7.373) | 31.870 (7.355) |
| Years of Experience<br>in Nursing Profession |  | 8.925 (7.483) | 7.941 (5.792) | 8.391 (6.633) |
| Years of Experience<br>in the Organization |  | 4.656 (4.965) | 5.581 (5.05) | 5.157 (5.030) |
| Gender | Male | 101 (28.77) | 61 (15.8) | 162 (22.04) |
|  | Female | 250 (71.30) | 324 (84.2) | 574 (78.10) |
| Marital Status | Married | 114 (32.48) | 198 (51.4) | 312 (42.45) |
|  | Unmarried | 237 (67.52) | 187 (48.6) | 424 (57.69) |
| Education | BSN | 276 (78.63) | 370 (96.1) | 646 (87.89) |
|  | Ma/PhD | 75 (21.37) | 15 (3.9) | 90 (12.24) |
| Employment | Fulltime | 301 (85.75) | 282 (73.2) | 583 (79.32) |
|  | Contracted | 50 (14.25) | 103 (26.8) | 153 (20.82) |
| Attendance in<br>COVID-19 related<br>trainings | Yes | 172 (49) | 191 (49.6) | 363 (49.39) |
|  | No | 179 (51) | 194 (50.4) | 373 (50.75) |
| Awareness of<br>COVID-19 protocol in<br>the workplace | Yes | 326 (92.88) | 374 (97.1) | 700 (95.24) |
|  | No | 25 (7.12) | 11 (2.9) | 36 (4.90) |
| Readiness to care for<br>COVID-19 patients | Unprepared | 15 (3.7) | 16 (4.2) | 28 (3.9) |
|  | Somewhat<br>unprepared | 41 (11.68) | 94 (24.2) | 128 (18) |
|  | Unsure | 153 (43.59) | 131 (34) | 278 (39.2) |
|  | Somewhat<br>prepared | 109 (31.05) | 127 (33) | 231 (32.5) |
|  | Prepared | 33 (9.40) | 17 (4.4) | 45 (6.3) |
| Willingness to care for<br>COVID-19 patients | Absolutely not | 35 (9.97) | 10 (2.6) | 41 (6.11) |
|  | Probably not | 31 (8.83) | 44 (11.4) | 69 (10.19) |
|  | Unsure | 123 (35.04) | 139 (36.1) | 255 (35.6) |
|  | Probably yes | 92 (26.21) | 150 (39) | 237 (32.88) |
|  | Absolutely yes | 70 (19.94) | 42 (10.9) | 108 (15.22) |
| COVID-19 Anxiety | < 8.9 | 221 (62.96) | 112 (29.091) | 333 (45.24) |
|  | > 9 | 130 (37.04) | 273 (70.91) | 402 (54.76) |

Overall, the prevalence rate of coronaphobia was 54.76% (*n* = 402): 37.04% (*n* = 130) in hospital nurses and 70.91% (*n* = 273) in public health nurses. Bivariate analysis using an independent *t*-test showed significantly higher scores on social support, personal resilience, and perceived general health measures among hospital nurses than public health nurses (all *p* < 0.001). No significant difference (*t* = 0.740, *p* = 0.460) was noted with regards to psychological distress between the two groups of frontline nurses **(Table 2)**.

**Table 2.** Comparison in job outcomes among nurses.

| Variables | Hospital Nurses |  | Public Health Nurses |  | t-test | p value |
| --- | --- | --- | --- | --- | --- | --- |
|  | Mean | SD | Mean | SD |  |  |
| COVID-19 Anxiety | 8.440 | 4.327 | 12.224 | 5.023 | -10.644* | 0.001 |
| Social Support | 3.956 | 0.761 | 3.219 | 0.791 | 12.579* | 0.001 |
| Personal Resilience | 4.190 | 0.688 | 3.219 | 0.963 | 15.204* | 0.001 |
| Perceived Health | 3.803 | 0.877 | 3.379 | 0.870 | 6.443* | 0.001 |

\*p < 0.001

Bivariate analyses showed a significant correlation between CAS and nurses’ age (*r* = 0.122, *p* = 0.001), years of experience in the organization (*r* = 0.088, *p* = 0.019), and readiness (r = −0.139, *p* = 0.001) and willingness (*r* = −0.153, *p* = 0.001) to manage infected patients. Further, social support, personal resilience, and perceived general health correlated significantly with CAS (all *p* < 0.001). Higher scores in the CAS were observed in female nurses (*t* = −4.522, *p* = 0.001), full-time nurses (*t* = −5.669, *p* = 0.001), those who were married (*t* = 4.925, *p* = 0.001), and those who held BSN degree (*t* = 2.239, *p* = 0.025) **(Table 3)**.

**Table 3.** Correlation between nurse characteristics and coronavirus anxiety.

| Characteristics | Category | Mean | SD | Test statistics | p value |
| --- | --- | --- | --- | --- | --- |
| Age<sup>†</sup> |  |  |  | 0.122 | 0.001 |
| Years of Experience in Nursing Profession<sup>†</sup> |  |  |  | 0.008 | 0.828 |
| Years of Experience in the Organization<sup>†</sup> |  |  |  | 0.088 | 0.019 |
| Readiness to care for COVID-19 patients<sup>†</sup> |  |  |  | -0.139 | 0.001 |
| Willingness to care for COVID-19 patients<sup>†</sup> |  |  |  | -0.153 | 0.001 |
| Social Support<sup>†</sup> |  |  |  | -0.271 | 0.001 |
| Personal Resilience<sup>†</sup> |  |  |  | -0.305 | 0.001 |
| General Health<sup>†</sup> |  |  |  | -0.163 | 0.001 |
| Gender<sup>‡</sup> | Male | 8.797 | 4.443 | -4.522 | 0.001 |
|  | Female | 10.915 | 5.137 |  |  |
| Marital Status<sup>‡</sup> | Married | 11.549 | 5.260 | 4.925 | 0.001 |
|  | Unmarried | 9.686 | 4.779 |  |  |
| Education<sup>‡</sup> | BSN | 10.633 | 5.137 | 2.239 | 0.025 |
|  | MS/PhD | 9.233 | 4.309 |  |  |
| Job Status<sup>‡</sup> | Fulltime | 10.005 | 5.064 | -5.669 | 0.001 |
|  | Contracted | 12.797 | 4.465 |  |  |
| Attendance in COVID-19 related trainings<sup>‡</sup> | Yes | 10.466 | 4.999 | -0.119 | 0.905 |
|  | No | 10.511 | 5.151 |  |  |
| Availability of COVID-19 protocol<sup>‡</sup> | Yes | 10.502 | 5.102 | 0.495 | 0.626 |
|  | No | 10.048 | 4.117 |  |  |

<sup>†</sup>Pearson r correlation
<sup>‡</sup>t-test for independent group
\*p < 0.05; \*\*p < 0.01; \*\*\*p < 0.001

Multiple linear regression identified nurses’ gender, marital status, job status, and personal resilience as predictors of COVID-19 anxiety **(Table 4)**. Female nurses (*β* = 0.146, *p* = 0.001) reported a higher level of anxiety than did male nurses. Further, part-time nurses (*β* = 0.132, *p* = 0.001) and those who were married (*β* = 0.101, *p* = 0.009) had higher anxiety scores than those who were unmarried and part-time nurses. Additionally, personal resilience (*β* = −0.154, *p* = 0.005) predicted dysfunctional anxiety related to coronavirus.

**Table 4.** Factors associated with coronavirus anxiety among nurses.

| Predictors | B | Std.<br>Error | $\beta$ | $t$ | Sig. | 95% Confidence Interval | |
| --- | --- | --- | --- | --- | --- | --- | --- |
|  |  |  |  |  |  | Lower<br>Bound | Upper<br>Bound |
| (Constant) | 12.921 | 1.601 |  | 8.069 | 0.001 | 9.777 | 16.065 |
| Age | 0.059 | 0.036 | 0.085 | 1.652 | 0.099 | -0.011 | 0.129 |
| Years in the organization | -0.034 | 0.050 | -<br>0.034 | -0.687 | 0.493 | -0.132 | 0.064 |
| Gender: (R: Male) |  |  |  |  |  |  |  |
| Female | 1.846 | 0.457 | 0.146 | 4.043 | 0.001 | 0.950 | 2.742 |
| Marital Status: (R: Unmarried) |  |  |  |  |  |  |  |
| Married | 1.034 | 0.395 | 0.101 | 2.621 | 0.009 | 0.259 | 1.809 |
| Education: (R: MSN/PhD) |  |  |  |  |  |  |  |
| BSN | 0.059 | 0.608 | 0.004 | 0.097 | 0.923 | -1.135 | 1.254 |
| Job Status: (R: Full Time) |  |  |  |  |  |  |  |
| Contracted | 1.774 | 0.485 | 0.132 | 3.659 | 0.001 | 0.822 | 2.726 |
| Readiness to care for COVID-19 patients | -0.285 | 0.196 | -<br>0.053 | -1.453 | 0.147 | -0.671 | 0.100 |
| Social Support | -0.521 | 0.315 | -<br>0.088 | -1.653 | 0.099 | -1.141 | 0.098 |
| Personal Resilience | -0.801 | 0.285 | -<br>0.154 | -2.815 | 0.005 | -1.361 | -0.242 |
| General Health | -0.198 | 0.214 | -<br>0.035 | -0.927 | 0.354 | -0.617 | 0.221 |

F = 13.801, ( $p < 0.001$ ), $R^2 = 16.8$

## Discussion

There is compelling evidence that nurses who are on the frontlines of combating coronavirus infection suffer from psychological, emotional, and metal consequences of the pandemic (Stelnicki *et al*., 2020; Leng *et al*., 2020). Surprisingly, most of these studies focus on the consequence of the pandemic on the mental and psychological health and well-being in frontline hospital nurses, while studies involving public health nurses remain elusive. Hence, this study was undertaken to compare the prevalence rate and associated factors of dysfunctional anxiety related to COVID-19 among frontline hospital and public health nurses.

About half of frontline nurses had no training related to coronavirus, while a smaller percentage reported to be absolutely willing (15.2%) and fully prepared (6.3%) to manage and care for coronavirus patients. This result is surprising considering the crucial role frontline hospital and public health nurses play in the achievement of the overall national healthcare plan and strategy to control, contain, and manage victims of the coronavirus pandemic. This result is an affirmation of the previous studies where nurses were reported to lack preparedness and willingness to manage patients afflicted with highly contagious diseases (e.g., H1N1, Ebola, COVID-19) (Baduge *et al*., 2018; McMullan *et al*., 2016; Labrague & De los Santos, 2020a). In the context of the coronavirus pandemic, an earlier study involving frontline hospital nurses in the Philippines showed that only less than 10% of nurses confirmed their complete readiness to care for confirmed cases of coronavirus, while only 20.3% were completely willing to handle infected patients. The need for COVID-19 related trainings and measures to improve nurses’ willingness and readiness was earlier highlighted in the study by Labrague and De los Santos (2020a), in which access to COVID-19 training to frontline nurses is raised to ensure that they are fully prepared and remain current with the different approaches in the management of patients affected by the pandemic. Earlier evidence has shown the importance of adequate training and capacity building in improving the willingness, readiness, and competence of nurses and other healthcare workers in responding to any disasters, emergency situations, and infectious disease outbreaks (Labrague *et al*., 2018; Labrague & De los Santos, 2020b).

Higher perceptions of social support, personal resilience, and general health among frontline nurses as compared to public health nurses is somewhat expected. This result may be related to the different work conditions in the two settings as hospital nurses often have the support of the physician and other healthcare team members in the care management of the patients, while public health nurses, in most cases, are solely responsible for the overall patient care and management (Dor *et al*., 2018). In addition, the hospital settings are often more equipped with the necessary supplies, equipment, and mechanisms necessary when handling and managing COVID-19 patients than in the community settings. These factors could possibly explain higher ratings on the social support, personal resilience, and general health measures among frontline hospital nurses.

Using the cut-off score set by Lee *et al*. (2020), the combined prevalence rate of coronaphobia was 55.6%, with 70.9% of public nurses and 37.8% of hospital nurses reporting coronaphobia. The increased percentage of public health nurses reporting dysfunctional anxiety levels related to coronavirus may be due to the fact that public health nurses, when compared to nurses working in the hospitals, are less equipped with the necessary supplies and equipment (e.g., PPE) to protect themselves from contracting the virus. This is particularly true in resource-scarce areas where resources and infrastructures are not made available. Further, nurses in the hospitals are generally aware of the diagnosis of the patients upon admission; hence, they can best protect themselves from exposure to the virus. Conversely, public health nurses have to deal with so many individuals in the community, and often, they are not fully aware of their complete history (e.g., travel history, contact with confirmed cases), making them vulnerable to or at risk for exposure to infected patients. Further, increased social support and higher personal resilience among hospital nurses may have contributed to lower levels of coronavirus anxiety when compared to public health nurses. Overall, the percentage of nurses reporting coronaphobia in the current study was higher when compared to previous studies where dysfunctional anxiety related to COVID-19 ranged from 3.3% to 46.3% (Chorwe-Sungani 2020; Saricam, 2020; Labrague & De los Santos, 2020a).

An important finding of the study was that gender predicted coronaphobia, with female nurses reporting a higher level of dysfunctional anxiety than male nurses. This result is rather expected as a result of gender differences when it comes to expression of emotions such as fear, anxiety, and sadness. Earlier studies have shown that women are more likely to express their emotions, while men tend to distract themselves away from these emotions (Chaplin *et al*., 2008; Tolin & Foa, 2008). Further, this gender disparity when it comes to reporting anxiety related to coronavirus mirrors the prominence of gender stereotypes within the Philippine culture that fear or anxiety is more acceptable for women than men. Nevertheless, this result provided support to previous studies in which female nurses were found to be more anxious than male nurses during the pandemic (Bitan *et al*., 2020; Pappa *et al*., 2020). Using a similar tool, Mora-Magan *et al*. (2020) found a similar pattern of relationship whereby female healthcare workers reported significantly higher scores in the CAS when compared to male nurses. Aside from anxiety, previous studies have also shown a higher tendency for female nurses to develop general anxiety disorder, depressive disorder, and post-traumatic stress symptoms during the pandemic than male nurses (Pouralizadeh *et al*., 2020; Han *et al*., 2020), suggesting the immediate need to address this concern through gender-tailored interventions.

Regression analysis identified marital status as a significant predictor of coronaphobia, with married nurses reporting a higher anxiety than unmarried nurses. This result may be attributed to increased fears of infecting their family or fear of harbouring the virus at work and bringing it home. The added family responsibilities and obligations may also have contributed to increased emotional reactions among married nurses. This result corroborates previous studies in which married nurses manifested higher levels of anxiety and fear related to coronavirus due to family reasons, including issues related to absence of childcare services during the mandatory lockdown (Shanafelt *et al*., 2020), fear of unknowingly infecting their loved ones (Saricam 2020), and fear of taking the virus home (Wu *et al*., 2009).

Results identified job status as a significant predictor of dysfunctional anxiety related to coronavirus, with contracted nurses experiencing higher levels of COVID-19 anxiety than full-time nurses. Caution should be exercised when interpreting this result due to the small number of contracted nurses included in the study. As contracted nurses typically fill in for staffing and may have no permanent unit of assignment, they are more likely than full-time nurses to experience anxiety as a result of lack of familiarity with the unit or ward processes, routines, and management of care for coronavirus patients. This result confirmed earlier research in which part-time nurses were seen to experience higher anxiety levels related to COVID-19 (Labrague & De los Santos, 2020b). Additional support for contracted nurses may be needed to sustain their mental and psychological health when caring for coronavirus patients.

Bivariate analysis also showed significant correlation between dysfunctional anxiety and nurses’ age and years of experience in the organisation; however, this pattern of relationship became insignificant when the multivariate analysis was conducted. Nevertheless, increased anxiety levels in older nurses who were in the organisation for a longer amount of time may be attributed to the increased perceived risk of contracting the virus, as mounting evidence has identified age as an important predictor of COVID-19 mortality and morbidity. In other words, as age increases, the risk for contracting the severe illness associated with COVID-19 and even for death is also increased (Applegate & Ouslander, 2020). Our result is different to that of Mora-Magana *et al*. (2020), where younger HCWs and those with less work experience tend to be anxious and depressed during the pandemic.

Personal resilience had a significant direct effect on nurses’ level of anxiety related to COVID-19. In other words, non-resilient nurses tend to experience more dysfunctional anxiety levels related to the virus than those nurses with higher levels of resilience. This result is rather expected as personal resilience has been found to support an individual when faced with adversity and distress, allowing them to successfully bounce back (Foster *et al*., 2020). This result is a confirmation of the earlier work conducted at the national level linking resilience with lower levels of anxiety among frontline hospital nurses due to pandemic (Labrague & De los Santos, 2020a). Further, this study result provided support to international studies identifying nurses’ personal resilience as a protective factor against a variety of mental and psychological issues, such as emotional exhaustion, depression, psychological distress, fatigue, and anxiety (Hu *et al*., 2020; Cooper *et al*., 2020). This result emphasised the relevance of implementing organisational measures to foster resilience in nurses and sustain their emotional and psychological health and well-being during the pandemic. As dysfunctional anxiety related to COVID-19 has been associated with job dissatisfaction, job stress, and higher turnover intention (Labrague & De los Santos, 2020b; Irshad *et al*., 2020), instituting measures to improve resilience in frontline nurses may be vital and should be prioritised by nursing and hospital administrators.

### Limitations of the Study

A few limitations of this study should be considered when analysing and interpreting the results. First, while sample calculation was conducted to determine the required sample size, increasing the sample size may be necessary to detect large effect size. Further, due to the nature of the research design, causality may be a challenge; hence, future studies using a more rigorous research design are recommended. While the coronavirus anxiety scale has been found valid and reliable to measure dysfunctional levels of anxiety, measures to clinically and accurately diagnose coronaphobia among nurses are imperative. Future studies should be undertaken considering other factors not included in the current study (e.g., self-efficacy, locus of control, personality) which could potentially affect the occurrence of dysfunctional anxiety levels.

## Conclusion

Coronaphobia is prevalent among frontline Filipino nurses, with more public health nurses experiencing coronaphobia than hospital nurses. Heightened anxiety related to coronavirus was commonly observed in female nurses, those who held contracted job status, and those who were married. As level of personal resilience predicted COVID-19 anxiety in nurses, interventions geared towards enhancing resilience in nurses through evidence-based education and training are essential to strengthen nurses’ defences against the emotional, mental, and psychological consequences of the pandemic. Future studies should focus on testing interventions to improve nurses’ resilience to effectively bounce back from adversity and effectively cope with stress caused by the coronavirus pandemic.

## Data Availability

Data is available upon request.

